# Associations of genetic and infectious risk factors with coronary heart disease

**DOI:** 10.1101/2022.04.13.22273812

**Authors:** Flavia Hodel, Zhi Ming Xu, Christian W. Thorball, Roxane de La Harpe, Prunelle Letang-Mathieu, Nicole Brenner, Julia Butt, Noemi Bender, Tim Waterboer, Pedro Marques-Vidal, Peter Vollenweider, Julien Vaucher, Jacques Fellay

## Abstract

**Background and Purpose:** Coronary heart disease (CHD) is one of the most pressing health problems of our time and a major cause of preventable death. CHD results from complex interactions between genetic and environmental factors. Using multiplex serological testing for persistent or frequently recurring infections and genome-wide analysis in a prospective population study, we delineate the respective and combined influences of genetic variation, infections, and low-grade inflammation on the risk of incident CHD.

**Participants and Methods:** Study participants are enrolled in the CoLaus|PsyCoLaus study, a longitudinal, population-based cohort with baseline assessments from 2003 through 2008 and follow-up visits every five years. We analyzed a subgroup of 3’459 individuals with available genome-wide genotyping data and immunoglobulin G levels for 22 persistent or frequently recurring pathogens. All reported CHD events were evaluated by a panel of specialists. We identified independent associations with incident CHD using univariable and multivariable stepwise Cox proportional hazards regression analyses.

**Results:** Of the 3’459 study participants, 210 (6.07%) had at least one CHD event during the 12 years of follow-up. Multivariable stepwise Cox regression analysis, adjusted for known cardiovascular risk factors, socioeconomic status and statin intake, revealed that high polygenic risk (hazard ratio (HR) 1.31, 95% CI 1.10–156, P = 2.64e-03) and infection with *Fusobacterium nucleatum* (HR 1.63, 95% CI 1.08–2.45, P = 1.99e-02) were independently associated with incident CHD.

**Conclusion:** In a prospective, population-based cohort, high polygenic risk and infection with *Fusobacterium nucleatum* have a small, yet independent impact on CHD risk.

## Introduction

Worldwide, cardiovascular diseases (CVDs) are the leading cause of mortality (1). An estimated 17.8 million people die from CVD each year, accounting for 32% of all deaths. CVD is a broad term for medical conditions involving the heart and blood vessels, such as coronary heart disease (CHD), congenital heart disease, cerebrovascular disease, peripheral arterial disease, rheumatic heart disease, deep vein thrombosis, and pulmonary embolism (2).

CHD is the most common type of heart disease (1). It is caused by atherosclerosis, a build-up of plaque inside the walls of the arteries that supply blood to the heart. CHD progresses over a long period of time and eventually evolves into symptoms such as chest pain (angina), tightness in the chest, breathing difficulties, and pain in the arms or shoulders (3). A complete blockage can cause a heart attack.

A combination of demographic, environmental and genetic factors contribute to the development of CHD (4, 5). The main risk factors associated with the development of CHD - smoking, diabetes, hyperlipidemia, and hypertension - have been established by extensive epidemiological research (6–9). Age is also an important risk factor for CHD (10). Finally, the incidence of CHD is greater in males than in females (10). Very recently, a new algorithm, named Systematic COronary Risk Evaluation 2 (SCORE2), was developed to predict the 10-year risk of first-onset CVD in European populations (11, 12). This score has replaced the existing HeartScore scoring system, and incorporates most of the risk factors mentioned above (13).

CHD also has an important genetic component. In 1938, the first familial risk model for CHD was described and later confirmed by clinical observations and large studies of twins and of longitudinal cohorts (14–17). Based on whole-genome approaches, the heritability of CHD has been estimated at 40-60%, even after controlling for known risk factors (18).

Multiple clinical studies have identified inflammatory risk factors that are predictive of future cardiovascular events (19–21). Endothelial dysfunction and subintimal cholesterol have been shown to trigger an inflammatory cascade, involving activated macrophages and leading to atherosclerotic lesions. At the molecular level, inflammasome formation in macrophages plays, through their production of interleukin (IL)-1*㬡*, an essential role in the propagation of inflammation. These cytokines are released, trigger various inflammatory cells, and produce IL-6 that in turn, stimulate C-reactive protein (CRP) production by the liver, which further enhances the inflammatory cascade within the vascular wall. Today, CRP is an established biomarker of systemic inflammation and a possible predictor of future cardiovascular events (20).

The recognition of atherosclerosis as an inflammatory disease has renewed interest in examining the role of pathogens in CHD and other CVDs. Nearly 150 years ago, acute infection with *Bacillus typhosus* (*B. typhosus*) was found to cause sclerosing changes in the arterial wall (22). A century later, the interest for a potential role of infection in atherosclerosis was renewed, with the discovery that CHD-positive individuals show an increased likelihood of having elevated levels of antibodies to *Chlamydia pneumoniae* (*C. pneumoniae*) (23). This was followed by the discovery of the association with CHD of several other infectious agents, including bacteria and viruses, such as *Helicobacter pylori* (*H. pylori*), hepatitis C virus (HCV), and human herpes viruses (24–27).

Although enormous progress has been made in the understanding of CHD pathogenesis, the overall picture of the combined contribution of infectious, inflammatory, and genetic factors to the risk of developing CHD in the general population remains incomplete. We here use data from the CoLaus|PsyCoLaus study, a well-characterized, longitudinal, population-based study from Switzerland, to obtain a more comprehensive view of the evidence for the respective contributions of these factors to CHD.

## Methods

### Study cohort

The CoLaus|PsyCoLaus study is a longitudinal population-based study initiated in Lausanne in 2003; it mainly investigates the biological, environmental, and genetic determinants of CVD (www.colaus-psycolaus.ch) (28). The study involves over 6’500 participants of European ancestry, who were recruited at random from the general population and represent approximately 10% sample of Lausanne citizens. Of the participants, 47.5% are men, and age at enrolment ranged from 35 to 75 years (mean ± standard deviation (SD): 51 ± 10.9). The study participants provided detailed phenotypic information through questionnaires, interviews, clinical and biological data. Nuclear deoxyribonucleic acid (DNA) was also extracted from the blood for whole-genome genotyping data. Every five years, follow-up interviews on the participants’ lifestyle and health status are conducted. There are three completed follow-ups and a fourth follow-up began in January 2022. The institutional Ethics Committee of the University of Lausanne, which later became the Ethics Commission of the Canton Vaud (www.cer-vd.ch), approved the CoLaus|PsyCoLaus study (reference 16/03, decisions of 13th January and 10th February 2003), and all participants gave written consent.

### Cardiovascular Phenotype

The medical records of the participants who reported a CHD event during their lifetime were collected and evaluated by an independent panel of specialists. Information on the cause of death was also collected prospectively during the study period. The full procedure was described previously (29). Only first events occurring after the baseline and up to day 4’500 after the baseline were included in the analysis, as only during this period were all participants reliably followed.

### DNA Genotyping Data and Polygenic Risk Score Calculation for Cardiovascular Phenotypes

The BB2 GSK-customized Affymetrix Axiom Biobank array was used to genotype DNA samples from 5’399 participants at approximately 800’000 single nucleotide polymorphisms (SNPs). After genotype imputation and quality control procedures, approximately 9 million SNPs were available for analysis (30). We then calculated, based on the risk effects of common SNPs, the CHD polygenic risk score (PRS) for each study participant. We used validated PRS from Inouye *et al*., available in the polygenic score catalog (31, 32). These scores and summary statistics were used to construct the CHD PRS (CHD-PRS) in our target cohort data by using the clumping and thresholding method of the PRSice-2 v2.2.7 software (33). A standardized method was used to obtain the PRS, by multiplying the risk allele dosage for each variant by the effect size and summing the scores across all selected variants. SNPs were clumped according to linkage disequilibrium (LD) (r2 < 0.1) within a 250 kb window.

### CHD Risk Evaluation

The risk of CHD for each participant was also assessed using the very recent SCORE2 and SCORE2-Older Persons (SCORE2-OP, for individuals >65 years of age) algorithms (11, 12). These two algorithms will be referred to as SCORE2 below. SCORE2 was derived, calibrated, and validated to predict the 10-year risk of first-onset CVD using data from 13 million individuals from >50 European prospective studies and national registries. To develop this algorithm, the authors used competing risk-adjusted and age- and sex-specific models including age, current smoking, systolic blood pressure, and total, low-density lipoprotein (LDL), and high-density lipoprotein (HDL) cholesterol. The authors also defined four risk regions in Europe on the basis of country-specific CVD mortality. For CoLaus|PsyCoLaus participants, calculations were based on the low-risk region corresponding to Switzerland. The raw scores of participants were standardized to Z-scores with approximately zero mean and unit variance before data analysis.

### Measurement of Inflammatory Biomarkers

Venous blood samples (50 mL) of the participants, in a fasted state, were drawn. Before cytokine assessment, the serum blood samples were stored at -80°C, then they were sent to the laboratory on dry ice. The measurements of high-sensitive CRP (hs-CRP), IL-1b, IL-6, and tumor necrosis factor *α* (TNF-*α*) cytokine levels were described previously in detail (34). Briefly, hs-CRP levels were assessed by immunoassay and latex HS (IMMULITE 1000–High, Diagnostic Products Corporation, LA, CA, USA). Cytokine levels were measured using a multiplexed particle-based flow cytometric cytokine assay on the flow cytometer (FC500 MPL, BeckmanCoulter, Nyon, Switzerland), thus following the manufacturer’s instructions. The lower limits of detection (LOD) for IL-1*㬡*, IL-6 and TNF-*α* were 0.2 pg/ml. Intra- and inter-assay coefficients of variation were, respectively, 15% and 16.7% for IL-1*㬡*, 16.9% and 16.1% for IL-6 and 12.5% and 13.5% for TNF-*α*. For quality control, repeat measurements were performed on 80 subjects randomly selected from the initial sample. Individuals with hs-CRP levels above 20 mg/L were assigned a value of 20 by the manufacturer therefore were removed from the hs-CRP analyses as indicative of acute inflammation.

### Serological Analyses

To assess the humoral responses to a total of 38 antigens derived from 22 persistent infectious agents, serum samples were analyzed by the Infections and Cancer Epidemiology Division at the German Cancer Research Center (Deutsches Krebsforschungszentrum, DKFZ) in Heidelberg (35, 36). Studied pathogens included 15 viruses (human polyomaviruses JC (JCV), BK (BKV), 6 (HPyV6), and WU (WUPyV), herpes simplex virus (HSV)-1, HSV-2, varicella zoster virus (VZV), Epstein–Barr virus (EBV), cytomegalovirus (CMV), human herpes virus 6A (HHV-6A), HHV-6B, HHV-7, Kaposi’s sarcoma-associated herpes virus (KSHV), parvovirus B19 (PVB-19), and rubella virus); six bacteria (*Chlamydia trachomatis* (*C. trachomatis*), *Clostridium tetani* (*C. tetani*), Cornybacterium diphteriae (C. diphteriae), *F. nucleatum, H. pylori*, and *Streptococcus gallolyticus* (*S. gallolyticus*)); and one parasite (*Toxoplasma gondii* (*T. gondii*)) (Supplementary Table 1). The seroreactivity was measured at a serum dilution of 1:1000 by using multiplex serology based on glutathione S-transferase (GST) fusion capture immunosorbent assays combined with fluorescent bead technology. For each infectious agent tested, the antibody responses were measured for 1 to 6 antigens and then expressed as a binary result (IgG positive or negative), based on the predefined median fluorescence intensity (MFI) thresholds. To define overall seropositivity against infectious agents when more than one antigen was used, we applied the pathogen-specific algorithms suggested by the manufacturer (see references in Supplementary Table 1).

### Statistical Analyses

Univariable and multivariable Cox proportional hazard models were used to explore the relationship between risk factors and CHD incidence in the CoLaus|PsyCoLaus study. Each variable was first screened in the univariable model. We then examined the proportional hazards assumption of the significant (P < 0.05) covariates by using the scaled Schoenfeld residuals. The residuals were plotted over time for each covariate to test for time-independence. Risk factors significantly associated with CHD in the univariable model were further evaluated using pairwise correlations. Finally, the identified risk factors were assessed using multivariable stepwise Cox regression analysis, adjusted for competing risk (i.e., SCORE2), socioeconomic status (i.e., gross monthly household income), and statin intake. Potential multicollinearity between statistically significant factors (P < 0.05) were identified using variance inflation factors (VIF). The existence of multicollinearity between co-variates was determined by a VIF value > 2. We performed all statistical analyses using R (version 4.1.1).

## Results

### Demographic and Serological Characteristics

A total of 3’459 CoLaus|PsyCoLaus participants with available phenotypic, serological, and genotypic data were included. Their characteristics are presented in Table 1.

**Table 1.**
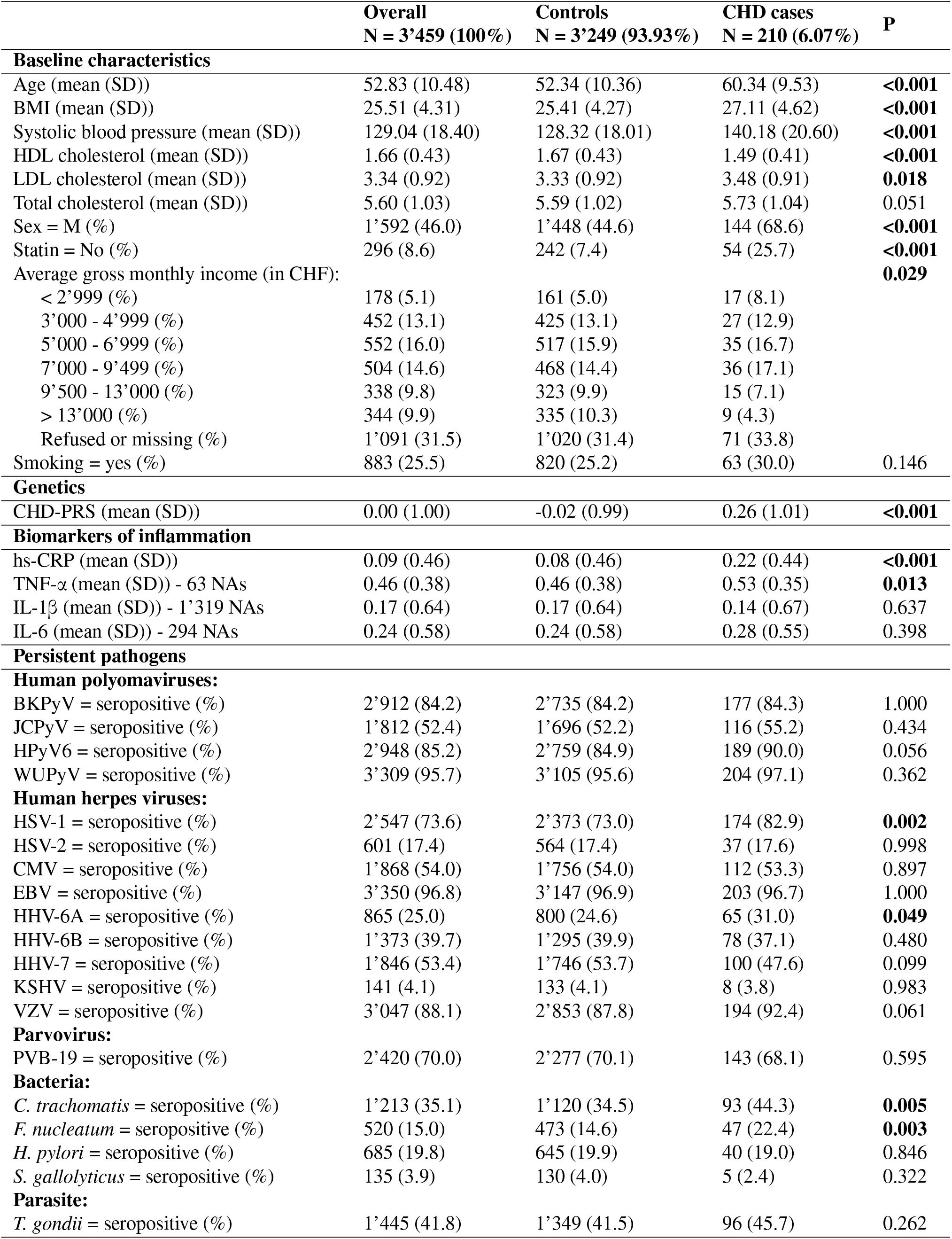
Baseline characteristics of 3’462 CoLaus|PsyCoLaus participants by CHD cases and controls. P-values are based on the t-test for continuous variables and Fisher’s exact test for categorical variables comparing the CHD Cases and Controls group.

|  | **Overall**<br>**N = 3'459 (100%)** | **Controls**<br>**N = 3'249 (93.93%)** | **CHD cases**<br>**N = 210 (6.07%)** | **P** |
| --- | --- | --- | --- | --- |
| **Baseline characteristics** |  |  |  |  |
| Age (mean (SD)) | 52.83 (10.48) | 52.34 (10.36) | 60.34 (9.53) | **<0.001** |
| BMI (mean (SD)) | 25.51 (4.31) | 25.41 (4.27) | 27.11 (4.62) | **<0.001** |
| Systolic blood pressure (mean (SD)) | 129.04 (18.40) | 128.32 (18.01) | 140.18 (20.60) | **<0.001** |
| HDL cholesterol (mean (SD)) | 1.66 (0.43) | 1.67 (0.43) | 1.49 (0.41) | **<0.001** |
| LDL cholesterol (mean (SD)) | 3.34 (0.92) | 3.33 (0.92) | 3.48 (0.91) | **0.018** |
| Total cholesterol (mean (SD)) | 5.60 (1.03) | 5.59 (1.02) | 5.73 (1.04) | 0.051 |
| Sex = M (%) | 1'592 (46.0) | 1'448 (44.6) | 144 (68.6) | **<0.001** |
| Statin = No (%) | 296 (8.6) | 242 (7.4) | 54 (25.7) | **<0.001** |
| Average gross monthly income (in CHF): |  |  |  | **0.029** |
| < 2'999 (%) | 178 (5.1) | 161 (5.0) | 17 (8.1) |  |
| 3'000 - 4'999 (%) | 452 (13.1) | 425 (13.1) | 27 (12.9) |  |
| 5'000 - 6'999 (%) | 552 (16.0) | 517 (15.9) | 35 (16.7) |  |
| 7'000 - 9'499 (%) | 504 (14.6) | 468 (14.4) | 36 (17.1) |  |
| 9'500 - 13'000 (%) | 338 (9.8) | 323 (9.9) | 15 (7.1) |  |
| > 13'000 (%) | 344 (9.9) | 335 (10.3) | 9 (4.3) |  |
| Refused or missing (%) | 1'091 (31.5) | 1'020 (31.4) | 71 (33.8) |  |
| Smoking = yes (%) | 883 (25.5) | 820 (25.2) | 63 (30.0) | 0.146 |
| **Genetics** |  |  |  |  |
| CHD-PRS (mean (SD)) | 0.00 (1.00) | -0.02 (0.99) | 0.26 (1.01) | **<0.001** |
| **Biomarkers of inflammation** |  |  |  |  |
| hs-CRP (mean (SD)) | 0.09 (0.46) | 0.08 (0.46) | 0.22 (0.44) | **<0.001** |
| TNF-α (mean (SD)) - 63 NAs | 0.46 (0.38) | 0.46 (0.38) | 0.53 (0.35) | **0.013** |
| IL-1β (mean (SD)) - 1'319 NAs | 0.17 (0.64) | 0.17 (0.64) | 0.14 (0.67) | 0.637 |
| IL-6 (mean (SD)) - 294 NAs | 0.24 (0.58) | 0.24 (0.58) | 0.28 (0.55) | 0.398 |
| **Persistent pathogens** |  |  |  |  |
| **Human polyomaviruses:** |  |  |  |  |
| BKPyV = seropositive (%) | 2'912 (84.2) | 2'735 (84.2) | 177 (84.3) | 1.000 |
| JCPyV = seropositive (%) | 1'812 (52.4) | 1'696 (52.2) | 116 (55.2) | 0.434 |
| HPyV6 = seropositive (%) | 2'948 (85.2) | 2'759 (84.9) | 189 (90.0) | 0.056 |
| WUPyV = seropositive (%) | 3'309 (95.7) | 3'105 (95.6) | 204 (97.1) | 0.362 |
| **Human herpes viruses:** |  |  |  |  |
| HSV-1 = seropositive (%) | 2'547 (73.6) | 2'373 (73.0) | 174 (82.9) | **0.002** |
| HSV-2 = seropositive (%) | 601 (17.4) | 564 (17.4) | 37 (17.6) | 0.998 |
| CMV = seropositive (%) | 1'868 (54.0) | 1'756 (54.0) | 112 (53.3) | 0.897 |
| EBV = seropositive (%) | 3'350 (96.8) | 3'147 (96.9) | 203 (96.7) | 1.000 |
| HHV-6A = seropositive (%) | 865 (25.0) | 800 (24.6) | 65 (31.0) | **0.049** |
| HHV-6B = seropositive (%) | 1'373 (39.7) | 1'295 (39.9) | 78 (37.1) | 0.480 |
| HHV-7 = seropositive (%) | 1'846 (53.4) | 1'746 (53.7) | 100 (47.6) | 0.099 |
| KSHV = seropositive (%) | 141 (4.1) | 133 (4.1) | 8 (3.8) | 0.983 |
| VZV = seropositive (%) | 3'047 (88.1) | 2'853 (87.8) | 194 (92.4) | 0.061 |
| **Parvovirus:** |  |  |  |  |
| PVB-19 = seropositive (%) | 2'420 (70.0) | 2'277 (70.1) | 143 (68.1) | 0.595 |
| **Bacteria:** |  |  |  |  |
| *C. trachomatis* = seropositive (%) | 1'213 (35.1) | 1'120 (34.5) | 93 (44.3) | **0.005** |
| *F. nucleatum* = seropositive (%) | 520 (15.0) | 473 (14.6) | 47 (22.4) | **0.003** |
| *H. pylori* = seropositive (%) | 685 (19.8) | 645 (19.9) | 40 (19.0) | 0.846 |
| *S. gallolyticus* = seropositive (%) | 135 (3.9) | 130 (4.0) | 5 (2.4) | 0.322 |
| **Parasite:** |  |  |  |  |
| *T. gondii* = seropositive (%) | 1'445 (41.8) | 1'349 (41.5) | 96 (45.7) | 0.262 |

During the follow-up of 4’500 days (12.3 years), at least one CHD event occurred in 210 individuals (6.07%). The number of participants with one, two, and three coronary events was 140, 47, and 14, respectively. Nine individuals had between 4 and 8 coronary events. Eligible study participants were on average 52.8 (SD ± 10.5) years of age at baseline, 54% were women and 25.5% were smokers. On average, their body mass index (BMI) was 25.5 (± 4.3) kg/m2, their systolic blood pressure was 129 (± 18) mmHg, and their HDL cholesterol level was 1.66 (± 0.43) mmol/L. The percentages of participants by average gross monthly income are 7.5 (<CHF 2’999), 19.1 (CHF 3’000-4’999), 23.3 (CHF 5’000-6’999), 21.3 (CHF 7’000-9’499), 14.3 (CHF 9’500-13’000) and 14.5% (CHF > 13’000).

For the measured biomarkers of inflammation, the log10-transformed mean (SD) values for hs-CRP, IL-1*β*, IL-6, TNF-*α* were 0.09 (± 0.46), 0.17 (± 0.64), 0.24 (± 0.58) and 0.46 (± 0.38), respectively.

We also investigated participants’ serostatus for the following 22 human pathogens: 15 viruses (BKV, JCV, HPyV6, WUPyV, HSV-1, HSV-2, VZV, EBV, CMV, HHV-6A, HHV-6B, HHV-7, KSHV, PVB-19, and rubella virus); six bacteria (*C. diphteriae, C. tetani, C. trachomatis, F. nucleatum, H. pylori*, and *S. gallolyticus*); and one parasite (*T. gondii*). The overall seropositivity ranged from 3.99% (*S. gallolyticus*) to 96.80% (EBV). The overall serostatus split between CHD positive (with at least one CHD event during follow-up) and CHD negative individuals, are shown in Figure 1. Rubella, *C. tetani* and *C. diphteriae* were excluded from further analyses as the antibodies detected against these pathogens were most likely induced by vaccination.

**Fig. 1.**
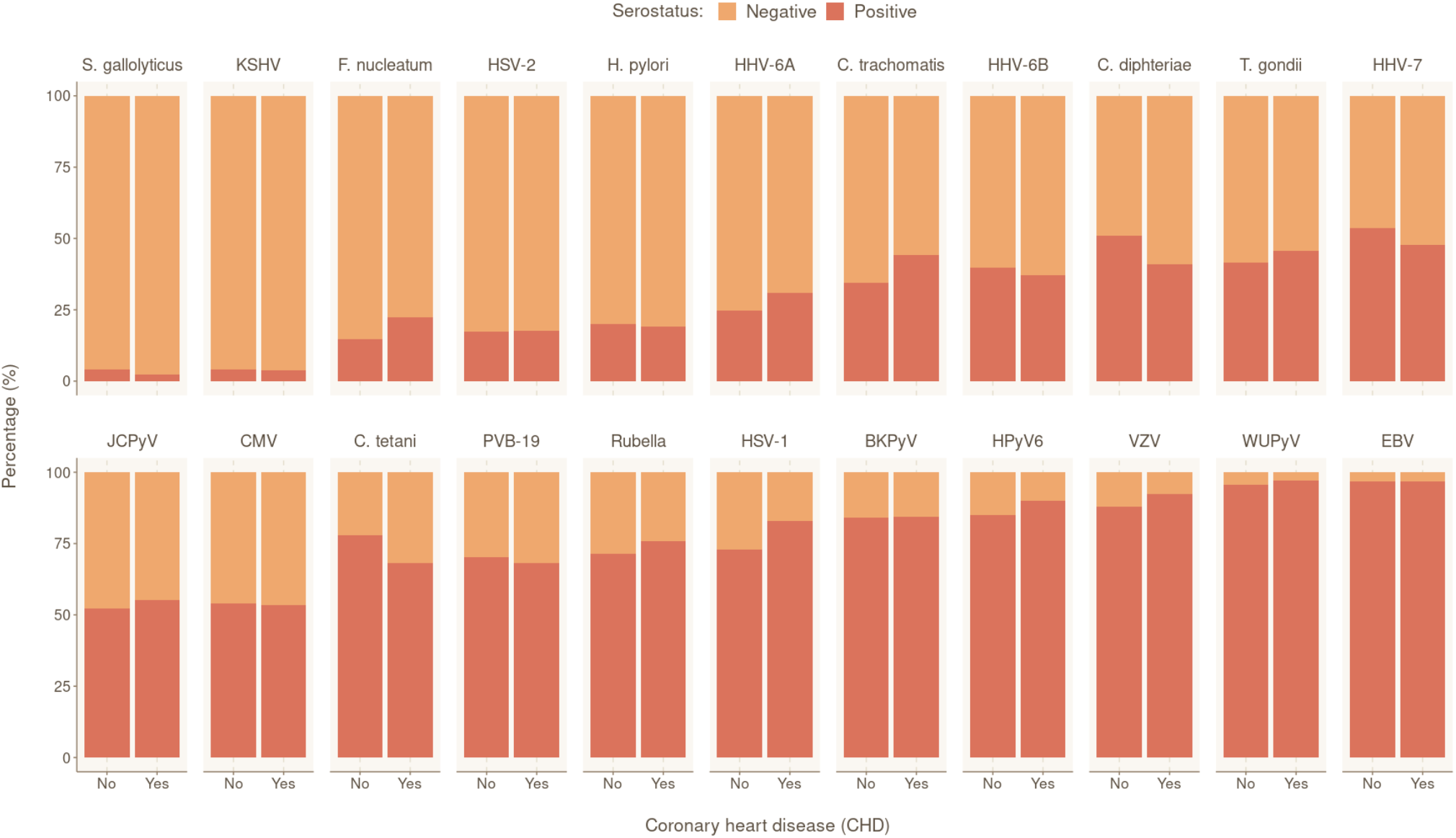
Prevalence of tested pathogens in CoLaus|PsyCoLaus study in participants with and without coronary heart disease. Overall serostatus for the 22 pathogens are shown in the CHD-positive group (indivdiuals with at least one CHD event during follow-up) or CHD-negative group. The y-axis indicates the relative percentage within each group. Pathogens are ranked in ascending order of overall seropositivity (all individuals combined).

### Univariable Predictors of CHD Incidence

To validate the utility of SCORE2 in our cohort, we tested its association with CHD. SCORE2 was significantly and positively associated with CHD (HR 1.72, 95% CI 1.61–1.85, P = 2.87e-61) (Suppl. Table 2). Finally, we observed a significant inverse association between average gross monthly income and CHD risk (HR 0.85, 95% CI 0.76–0.96, P = 7.27e-03).

To investigate the relationship between CHD and humoral response to infectious agents, we tested the association of serostatus for each of the included 19 persistent or frequently occurring pathogens with CHD. We found significant positive associations for six of them, including three herpes viruses, namely HSV-1 (HR 1.88, 95% CI 1.30–2.68, P = 6.52e-04), HHV-6A (HR 1.39, 95% CI 1.03–1.86, P = 2.89e-02), and VZV (HR 1.70, 95% CI 1.02–2.82, P = 4.25e02), one polyomavirus, HPyV6 (HR 1.66, 95% CI 1.06–2.61, P = 2.74e-02), and two bacteria, *F. nucleatum* (HR 1.66, 95% CI 1.20–2.29, P = 2.32e-02), and *C. trachomatis* (HR 1.45, 95% CI 1.11–1.91, P = 7.22e-03) (Supplementary Table 2).

To evaluate the impact of the biomarkers of inflammation on CHD risk, we tested the association of log10transformed levels of hs-CRP, IL-1b, IL-6 and TNF-a with CHD. We observed a positive relationship between individuals’ hs-CRP (HR 1.91, 95% CI 1.42–2.55, P = 1.51e-05) and TNF-a (HR 1.43, 95% CI 1.05–1.96, P = 3.08e-03) levels, and increased risk of CHD event (Supplementary Table 2).

Finally, we calculated a CHD-PRS for each subject to investigate the effect of common human genetic variations on CHD. As expected, we observed a significant association between the PRS and CHD (HR 1.32, 95% CI 1.16–1.51, P = 4.29e-05), confirming that genetic predisposition to CHD can be captured through CHD-PRS. To identify potential confounding due to population structure, the top three genetic principal components (PC1, PC2 and PC3) were also tested for association with CHD. All three principal components were not significantly associated with CHD (Supplementary Table 2).

### Co-linearity and Proportional Hazard Assumption Testing

We calculated pairwise correlations between all variables that were found to be significant in univariable analysis. Supplementary Figure 1 and Supplementary Figure 2 illustrate that no strong correlations exist between significant variables. The strongest correlation was observed between SCORE2 and average gross monthly income, and between seropositivity to HSV-1 and *C. trachomatis*, with Pearson’s correlation coefficient of -0.24 and 0.08, respectively. The proportionality assumption was tested for all significant variables using the Schoenfeld residuals. The residual tests indicated that all variables satisfied the proportional hazards assumption, revealing that the effect of all covariates are constant in time (Supplementary Figure 3). Finally, we also assessed potential co-linearity issues among predictors that could affect model fitting. No VIF value was indicative of co-linearity.

### Multivariable Model

To identify the independent risk factors of CHD in our cohort, we performed backward stepwise selection using a multivariable Cox proportional hazards model, starting with all the significant factors from the univariable models. The final multivariable analysis confirmed that SCORE2 (HR 1.96 per SD increase, 95% CI 1.74–2.22, P = 2.42e-27) is an independent prognostic factor of CHD (Figure 2). We also observed significant independent associations for statin intake (HR 2.24, 95% CI 1.50–3.35, P = 9.17e-05) and for seropositivity to *F. nucleatum* infection (HR 1.63, 95% CI 1.08–2.45, P = 1.99e-02). Comparing individuals who had a least one CHD event (CHD group) against those who had no event during the follow-up period (control group), 22.4% (47/210) of the individuals in the CHD group were seropositive to *F. nucleatum*, versus 14.6% (473/3’249) in the control group (P = 0.003) (Figure 1, Table 1). Lastly, we also observed a significant association between CHD occurrence and elevated CHD-PRS with a HR of 1.40 (95% CI 1.21–1.61, P = 3.32e-06) per SD increase.

To assess if the overall burden of infections contributed to increased risk of CHD, study participants were stratified according to their overall seropositivity index for measured pathogens, calculated by summing the number of pathogens for which they show seropositivity (range: 0-16). The numbers of individuals in each pathogen burden stratum are shown in Supplementary Figure 4. In the univariable Cox model, pathogen burden significantly increased the risk of CHD occurrence (HR 1.11, 95% CI 1.03–1.18, P = 9.59e-06). However, after adjustment with multivariable Cox proportional hazards regression, pathogen burden did not meet the level of significance for staying in the model.

**Fig. 2.**
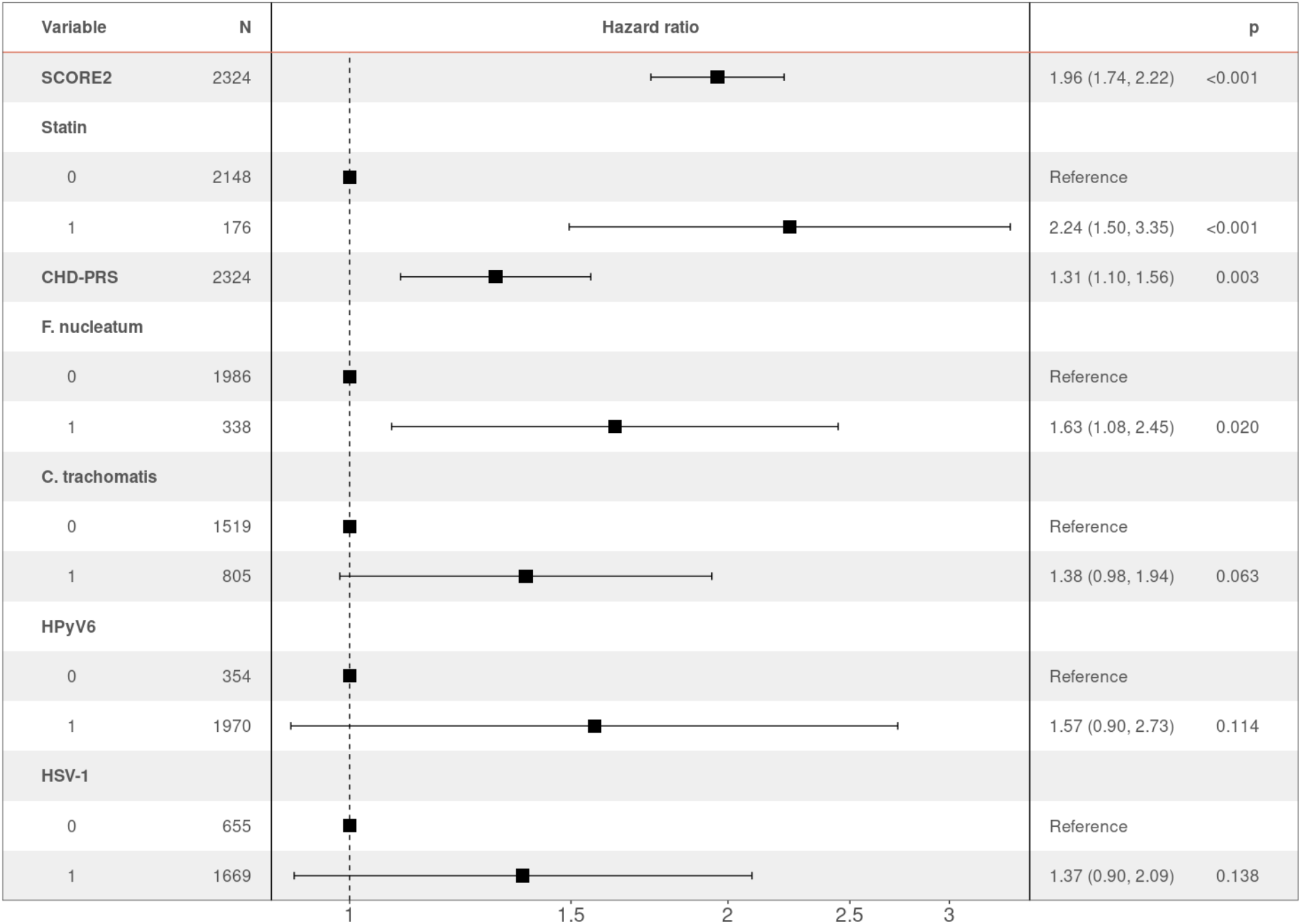
Hazard ratio (HR) and 95% confidence intervals of CHD occurrence according to associated factors. HR > 1 indicates an increased risk of CHD, whereas HR < 1 indicates a protective effect. Pvalues (p) for each factor based on the multivariable cox-regression are shown.

## Discussion

CHD is a complex disease that is influenced by demographic, environmental and genetic factors (4, 5). Infections have also been suspected to increase the risk of CHD, directly or through the induction of chronic inflammation (37). The present study investigated the independent and combined effects of these risk factors as possible prognostic indicators for the occurrence of CHD. We performed an event-free survival analysis of incident CHD using data from a longitudinal, population-based study, in which more than 6% of participants developed CHD over a 12-year study period.

We confirmed the utility of SCORE2 to predict CHD risk in our cohort (11). Of note, chronic inflammation reflected in hs-CRP level did not appear as an independent predictor of CHD in our analyses, as the univariable association signal was suppressed after adjustment for SCORE2 levels.

We studied the effect of human genetic determinants on CHD occurrence using PRS, and we reproduced previously observed effects: participants with a higher CHD-PRS have a greater risk of CHD, even after adjustment for all known factors (38, 39). This result confirms the existence of genetic susceptibility loci for CHD, and that the individual genetic background modulates CHD risk independently from age, sex or co-morbidities. Our work confirms the potential interest in using PRS to improve the prediction of coronary events.

We also evaluated the potential contribution of multiple persistent or frequently recurring pathogens to CHD after controlling for conventional CHD risk factors, socioeconomic status, and human genetic variability. We observed an association of CHD with detection of antibodies against *F. nucleatum*. This pathogen is very prevalent in humans (40– 42). *F. nucleatum* is an anaerobic bacterium that belongs to the normal flora of the oral cavity and plays an important role in the development and progression of oral diseases, such as gingivitis (gum inflammation) and periodontitis (infection of the gums). Under pathological conditions, the pathogen can spread by the hematogenous route to extra-oral systemic sites, including the gut and the female genital tract (43, 44). Studies have also suggested the involvement of *F. nucleatum* in CVD. First, by its capacity to directly migrate into arterial plaques, thus exacerbating atherosclerosis, and more recently, through the association of periodontitis and CVD (45– 50). Finally, it has been shown that periodontal pathogens are able to spread through the bloodstream from the buccal cavity to the arteries in patients with detectable coronary calcium, a very specific marker of atherosclerosis (51). In summary, the relationship between oral inflammations and CVD could be explained by the colonization of arterial walls and atherosclerosis plaques by dental bacteria, as well as by increased systemic inflammation due to oral infection. However, to date, no direct causality has been established.

HSV-1, HHV-6A, VZV, HPyV6, and *C. trachomatis* serologies, as well as total burden of infection, were associated with CHD occurrence in univariable models. However, these factors were not significantly associated in the multivariable analysis, suggesting that at least some of them could be indirect markers of socio-economic status.

Our data do not support the existence of the previously identified associations between CHD and *H. pylori*, or CMV. The conflicting reports of possible associations between these pathogens and CHD could be due to sample size but remain questionable. Further extensive, and high-quality studies are needed to thoroughly examine these associations and provide firm conclusions.

Our study has several limitations. As is the case for most longitudinal studies, the absence of data on individuals who dropped out before the end of the follow-up implies that some CHD events could have gone undetected. Also, the demographic information, as well as the clinical and laboratory measurements, were obtained at baseline, and we do not know whether participant information changed over time. Adjustment for risk factors measured at baseline does not account for clinical or demographic changes that could influence CHD outcomes. Similarly, we do not know how the antibody responses against the various antigens evolved over the 12 years of the study. We were also unable to replicate previously published observations of associations of CHD with *C. pneumoniae* and HCV as serologies for these pathogens were not available. From a more practical point of view, the identified association with *F. nucleatum* needs to be replicated and validated in independent cohorts and different populations. Finally, the clinical utility of including genetic and infection biomarkers in CHD prediction algorithms will need to be demonstrated.

## Conclusion

CHD is a multicomponent disease that is caused by demographic, environmental and genetic factors. Inflammation, possibly caused by persistent or frequently recurring infections, can contribute to its development. We identified a statistically significant association between the incidence of CHD and *F. nucleatum* infection, after adjustment for all established risk factors. We also confirmed that the individual polygenic risk of cardiovascular disease, calculated from genome-wide genotypes, represents an independent risk factor for incident CHD. Our results can help to better identify subjects at high risk for CHD and provide a rationale for future anti-infective prevention trials.

## Data Availability

The CoLaus|PsyCoLaus cohort data used in this study cannot be fully shared as they contain potentially sensitive patient information. As discussed with the competent authority, the Research Ethic Committee of the Canton of Vaud, transferring or directly sharing this data would be a violation of the Swiss legislation aiming to protect the personal rights of participants. Non-identifiable, individual-level data are available for interested researchers, who meet the criteria for access to confidential data sharing, from the CoLaus|PsyCoLaus Datacenter (CHUV, Lausanne, Switzerland). Instructions for gaining access to the CoLaus|PsyCoLaus data used in this study are available at (www.colaus-psycolaus.ch/professionals/how-to-collaborate/).

## ACKNOWLEDGEMENTS

We thank the participants in the CoLaus|PsyCoLaus study for their time and contribution to this study. We also thank all the clinical, academic, and administrative collaborators who helped with participant recruitment, study coordination, data collection, and storage.

## AUTHOR CONTRIBUTIONS

FH: Conceptualization, Methodology, Software, Formal analysis, Visualization, Writing – Original Draft. ZMX: Conceptualization, Methodology. CWT: Methodology, Resources. RdlH: Resources. PLM: Formal analysis. NBr: Resources. JB: Resources. NBe: Resources. TW: Investigation, Resources. PMV: Resources, Data Curation, Investigation. PV: Investigation. JV: Methodology, Resources. JF: Funding Acquisition, Project administration, Supervision, Writing – Original Draft. All co-authors reviewed the manuscript.

## CONFLICTS OF INTEREST

The authors have no related financial or non-financial interests to disclose.

## FUNDING

This project was supported by the Swiss National Science Foundation (grant 31003A_175603 to JF). The CoLaus|PsyCoLaus study was and is supported by research grants from GlaxoSmithKline, the Faculty of Biology and Medicine of Lausanne, and the Swiss National Science Foundation (grants 3200B0_105993, 3200B0_118308, 33CSCO_122661, 33CS30_139468, 33CS30_148401 and 33CS30_177535/1).

## DATA ACCESS

The CoLaus|PsyCoLaus cohort data used in this study cannot be fully shared as they contain potentially sensitive patient information. As discussed with the competent authority, the Research Ethic Committee of the Canton of Vaud, transferring or directly sharing this data would be a violation of the Swiss legislation aiming to protect the personal rights of participants. Non-identifiable, individual-level data are available for interested researchers, who meet the criteria for access to confidential data sharing, from the CoLaus|PsyCoLaus Datacenter (CHUV, Lausanne, Switzerland). Instructions for gaining access to the CoLaus|PsyCoLaus data used in this study are available at (www.colaus-psycolaus.ch/professionals/how-tocollaborate/).

## Notes

### Competing Interest Statement

The authors have declared no competing interest.

### Author Declarations

The institutional Ethics Committee of the University of Lausanne, which later became the Ethics Commission of the Canton Vaud (www.cer-vd.ch), gave ethical approval for the CoLaus|PsyCoLaus study (reference 16/03, decisions of 13th January and 10th February 2003), and all participants gave written consent.

