## Supplementary Material for "Associations of genetic and infectious risk factors with coronary heart disease"

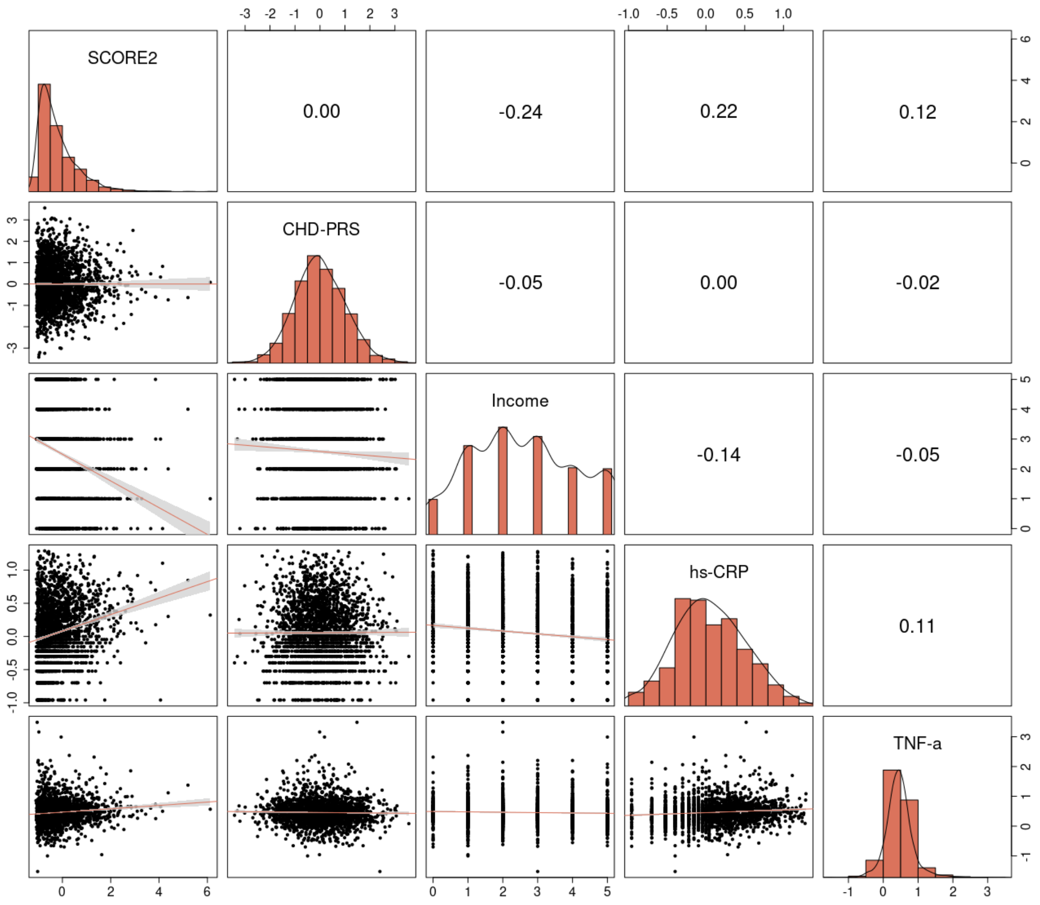


**Supplementary Figure 1. Pairwise correlations between quantitative characteristics significantly associated with CHD risk in the univariable Cox proportional hazard models.** Pearson's correlation values are displayed, along with linear fits between variables.

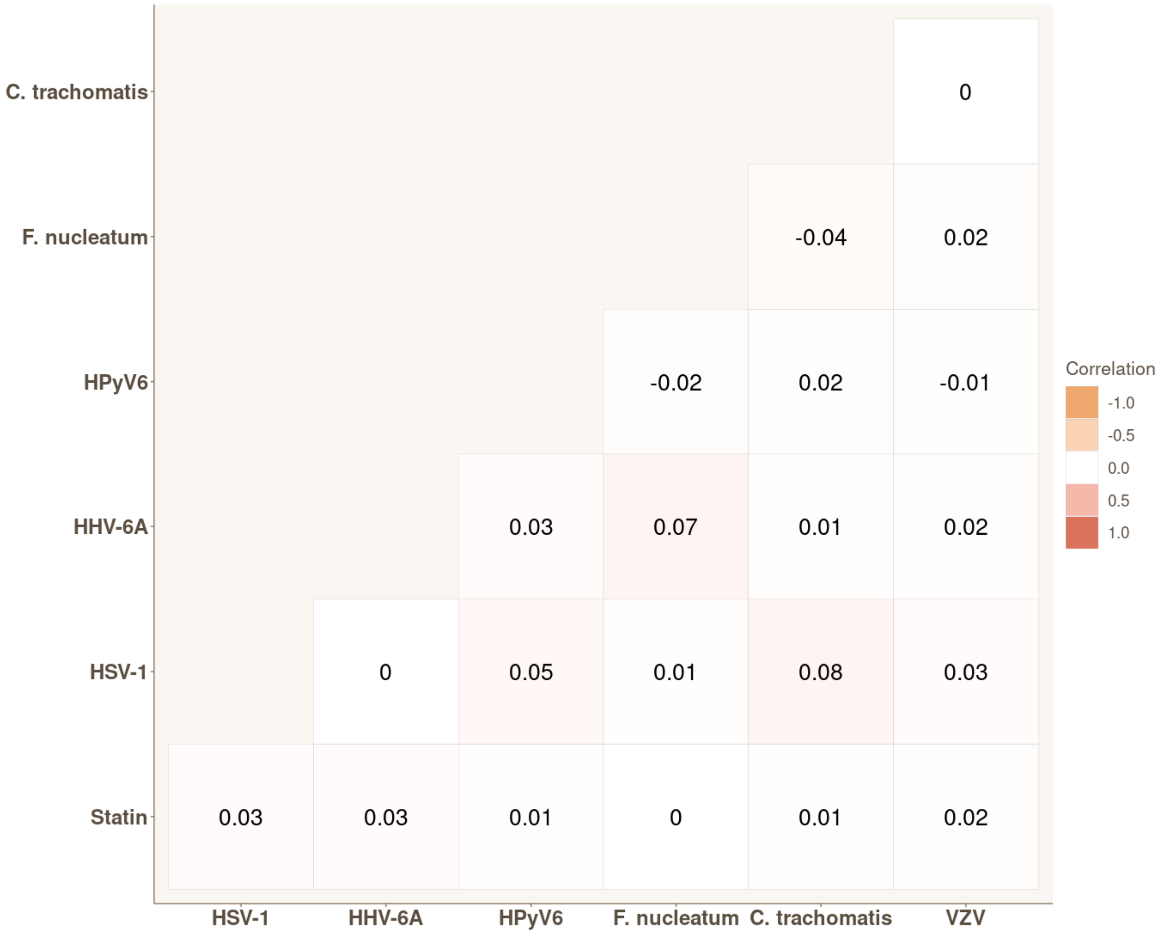


**Supplementary Figure 2. Pairwise correlations between binary variables significantly associated with CHD risk in the univariable Cox proportional hazard models.** Pearson's correlation values are displayed.

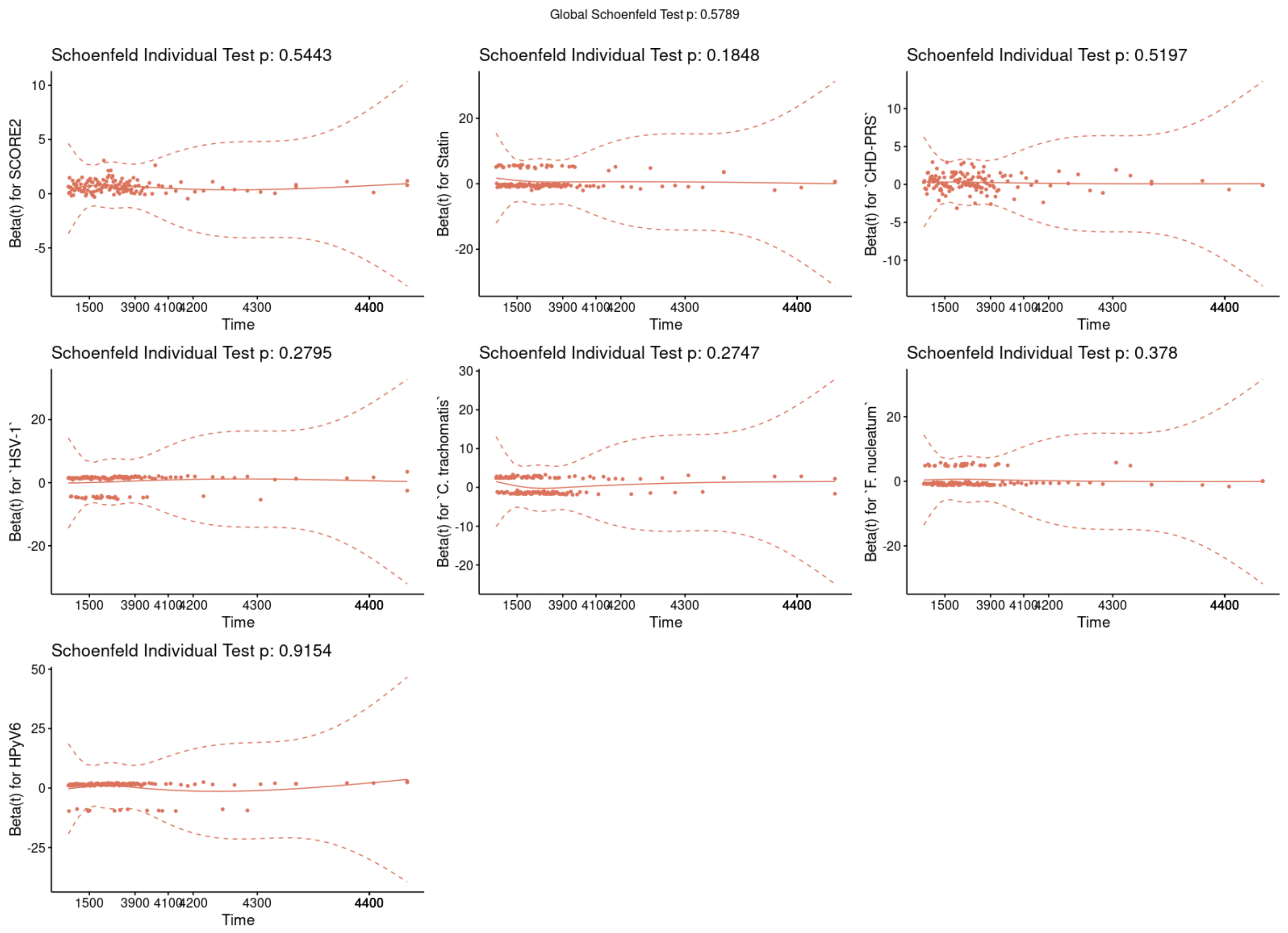


**Supplementary Figure 3. Graphical test of Proportional hazards assumption (Schoenfeld test).** The graphs show the scaled Schoenfeld residuals over time. The P-values (p) of the variables and the model as a whole were shown in the plot. A significant P-value (< 0.05) indicates that the variable violates the proportional-hazard assumption. The solid line represents the smoothing fitted spline, and the dashed lines the confidence bands at two standard errors. Global Schoenfeld Test P = 0.58.

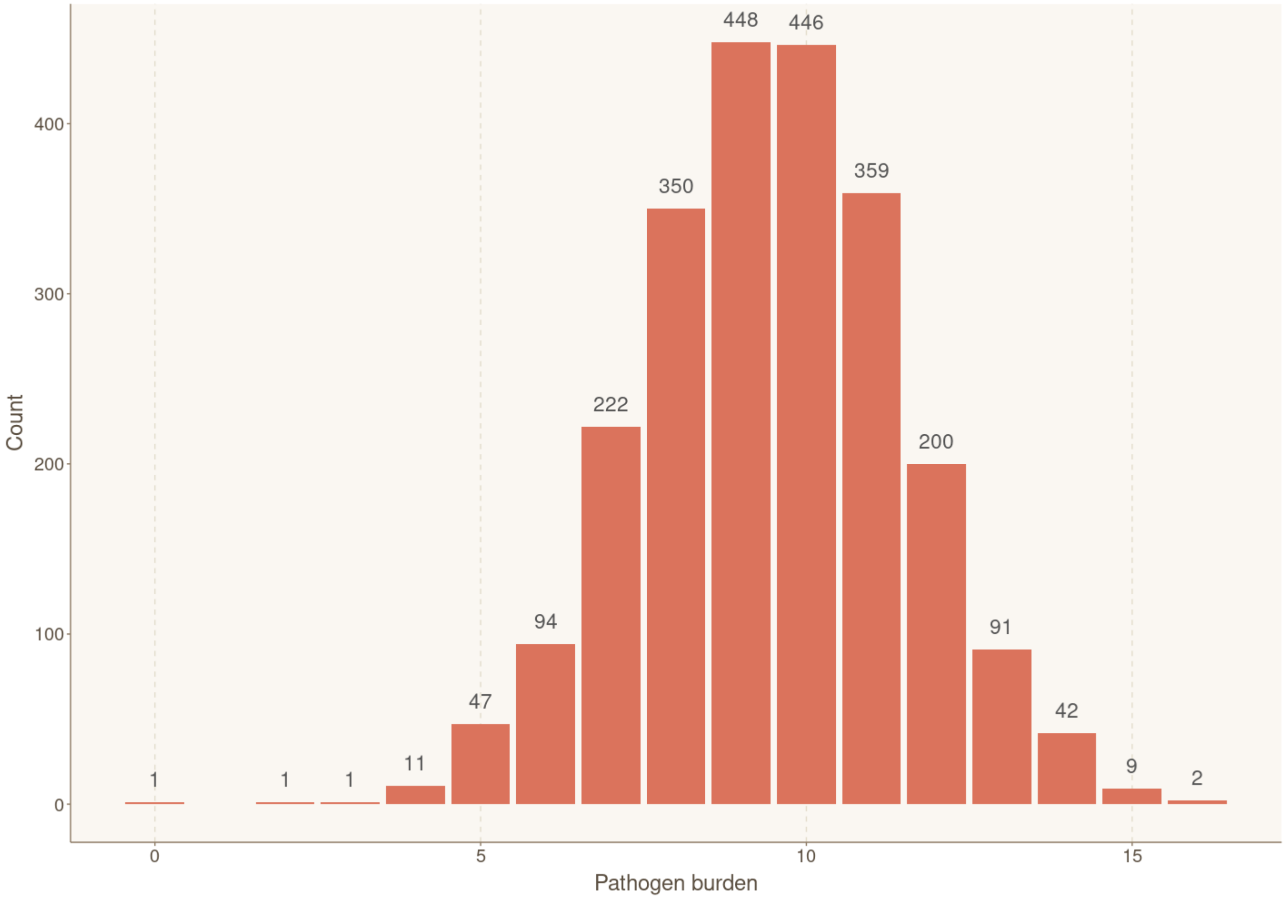


**Supplementary Figure 4. Distribution of individuals according to their exposure to infectious agents (pathogen burden).** Bar plot showing the number of participants for each cumulative number of positive serological results, reflecting simultaneous ongoing chronic/latent infections. Sample sizes for each group are shown above the box.

**Supplementary Table 1.** Characteristics of infectious agent specific antigens used on the Multiplex Serology platform in CoLaus|PsyCoLaus.

| Family | Pathogen | Antigen | (Predicted) function | Def. of seropositivity is based on | Reference |
|---|---|---|---|---|---|
| Human Polyomaviruses | BKV | VP1 | Major capsid protein | NA | (1–3) |
| | JCV | VP1 | Major capsid protein | NA | (1–3) |
| | HPyV6 | VP1 | Major capsid protein | NA | (1–3) |
| | WUPyV | VP1 | Major capsid protein | NA | (1–3) |
| Human herpes viruses | CMV | pp150<br>pp52<br>pp28 | Tegument protein<br>DNA binding protein<br>Capsid protein | At least 2 positive | (4) |
| | EBV | ZEBRA<br>EA-D<br>VCA p18<br>EBNA1 | Replication activator<br>Replication (polymerase accessory subunit)<br>Capsid protein<br>Replication, latent viral infection | At least 2 positive | (4) |
| | HHV-6 | IE1B<br>IE1A<br>p101K<br>p100 | Potential transactivator<br>Potential transactivator<br>Potential tegument protein<br>Potential tegument protein | Any HHV-6 = at least 1 positive<br>HHV-6A = IE1A and/or p100<br>HHV-6B = IE1B and/or p101K | (5–7) |
| | HHV-7 | U14 | Potential tegument protein | NA | Validation ongoing |
| | HSV-1 | gG | Membrane glycoprotein | NA | (4) |
| | HSV-2 | mgG | Membrane glycoprotein | NA | (4) |
| | KSHV | LANA3<br>K8.1 | Replication and long-term persistence<br>Structural glycoprotein | At least 1 positive | Validation ongoing |
| | VZV | gE/gI | Envelope glycoprotein | NA | (4) |
| Parvovirus | B19 | VP1unique | Minor capsid protein | NA | (8) |
| Rubella virus * | RV | E1 | Class II viral fusion protein | NA | (8) |
| *Chlamydia trachomatis* | Ct | pGP3 | Virulence factor | NA | (9) |
| *Clostridium tetani* * | Ct | TetX | Toxoid (heavy chain) | NA | (8) |
| *Corynebacterium diphtheriae* * | Cd | DTA | Toxoid (intracellular) | NA | (8) |
| *Fusobacterium nucleatum* | Fn | Fn0264<br>Fn1449<br>Fn1859 | Adhesin (FadA)<br>Type Va secretion system (Fap2)<br>Porin (FomA) | At least 1 positive (experimental) | (10) |
| *Helicobacter pylori* | Hp | HP 10 GroEL<br>HP 73 UreaseA<br>HP 547 CagA<br>HP 875 Catalase<br>HP 887 VacA<br>HP 1564 OMP | Chaperonin<br>Urease alpha subunit<br>Pathogenesis<br>Detoxification<br>Pathogenesis<br>Cell envelope | At least 3 positive | (11) |
| *Streptococcus gallolyticus* | Sg | Gallo2178 | *Pil1 pilus subunit (major pilin)* | NA | (12, 13) |
| *Toxoplasma gondii* | Tg | p22<br>sag-1 | Surface protein<br>Surface protein | At least 1 positive | (14) |

* Pathogens not taken forward due to lack of vaccination history in CoLaus|PsyCoLaus and/or the difficulty in identifying target antigens to ensure specificity of the test.

**Supplementary Table 2. Association of risk factors with CHD based on the univariable Cox proportional hazard analyses.**

| Variable | HR* (95% CI*) | P |
|---|---|---|
| **Baseline characteristics** | | |
| SCORE2 | 1.72 (1.61 – 1.85) | **2.87e-61** |
| Statin | 3.82 (2.80 – 5.22) | **3.13e-17** |
| Average gross monthly income | 0.85 (0.76 – 0.96) | **7.27e-03** |
| **Genetics** | | |
| CHD-PRS | 1.32 (1.16 – 1.51) | **4.29e-05** |
| PC1 | 74-28 (0.03 – 195096) | 2.83e-01 |
| PC2 | 0.12 (00.0 – 728) | 6.37e-01 |
| PC3 | 0.33 (0.00 – 1131) | 7.88e-01 |
| **Biomarkers of inflammation** | | |
| hs-CRP** | 1.91 (1.42 – 2.55) | **1.51e-05** |
| TNF-α** | 1.43 (1.05 – 1.96) | **2.46e-02** |
| IL-1β** | 0.93 (0.70 – 1.25) | 6.40e-01 |
| IL-6** | 1.10 (0.88 – 1.37) | 4.21e-01 |
| **Human polyomaviruses** | | |
| BKPyV | 1.05 (0.72 – 1.52) | 7.97e-01 |
| JCPyV | 1.14 (0.87 – 1.50) | 3.45e-01 |
| HPyV6 | 1.66 (1.06 – 2.61) | **2.74e-02** |
| WUPyV | 1.45 (0.65 – 3.27) | 3.67e-01 |
| **Human herpes viruses** | | |
| HSV-1 | 1.88 (1.30 – 2.68) | **6.52e-04** |
| HSV-2 | 1.05 (0.74 – 1.50) | 7.82e-01 |
| CMV | 1.00 (0.76 – 1.31) | 9.85e-01 |
| EBV | 0.97 (0.46 – 2.06) | 9.38e-01 |
| HHV-6A | 1.39 (1.03 – 1.86) | **2.89e-02** |
| HHV-6B | 0.93 (0.70 – 1.23) | 5.90e-01 |
| HHV-7 | 0.79 (0.60 – 1.03) | 8.33e-02 |
| KSHV | 0.89 (0.44 – 1.80) | 7.39e-01 |
| VZV | 1.70 (1.02 – 2.82) | **4.25e-02** |
| **Parvovirus** | | |
| PVB-19 | 0.90 (0.68 – 1.21) | 4.93e-01 |
| **Bacteria** | | . |
| *C. trachomatis* | 1.45 (1.11 – 1.91) | **7.22e-03** |
| *F. nucleatum* | 1.66 (1.20 – 2.29) | **2.32e-03** |
| *H. pylori* | 0.95 (0.67 – 1.34) | 7.81e-01 |
| *S. gallolyticus* | 0.62 (0.26 – 1.51) | 2.92e-01 |
| **Parasite:** | | |
| *T. gondii* | 1.17 (0.90 – 1.54) | 2.45e-01 |
| **Pathogen burden** | 1.11 (1.03 – 1.18) | **3.25e-03** |

* HR = Hazard Ratio, CI = Confidence Interval
** log10-transformed
